# Brief Analysis of the ARIMA model on the COVID-19 in Italy

**DOI:** 10.1101/2020.04.08.20058636

**Authors:** Guorong Ding, Xinru Li, Fan Jiao, Yang Shen

## Abstract

Coronavirus disease 2019 (COVID-19) has been considered as a global threat infectious disease, and various mathematical models are being used to conduct multiple studies to analyze and predict the evolution of this epidemic. We statistically analyze the epidemic data from February 24 to March 30, 2020 in Italy, and proposes a simple time series analysis model based on the Auto Regressive Integrated Moving Average (ARIMA). The cumulative number of newly diagnosed and newly diagnosed patients in Italy is preprocessed and can be used to predict the spread of the Italian COVID-19 epidemic. The conclusion is that an inflection point is expected to occur in Italy in early April, and some reliable points are put forward for the inflection point of the epidemic: strengthen regional isolation and protection, do a good job of personal hygiene, and quickly treat the team leaders existing medical forces. It is hoped that the “City Closure” decree issued by the Italian government will go in the right direction, because this is the only way to curb the epidemic.

## 1. Introduction

Coronavirus disease 2019 (COVID-19) has been regarded as a global threat, which has attracted much attention since 2019. At present, the proliferation trend has been contained in China, and to prevent foreign import is the key point to evaluate whether we achieve the final success. However, Italy, which located in Europe, is in a serious stage of epidemic spread. Fundamentally, due to the long incubation period of the virus, the difficulty in identifying symptoms, negligence in the prevention and control of returnees, and the relatively low number of sick people, Italy and the whole of Europe have not been highly vigilant, which has brought an opportunity for the spread of the virus ^[1]^.

Various mathematical models are being used to conduct a variety of studies to analyze and predict the evolution of this epidemic. Reference [2] based on the SEIR kinetic model, taking into account the propagation mechanism, infection rate, and isolation measures of COVID-19, established a SEIR^+CAQ^ propagation kinetic model, which can be used to predict the trend of COVID-19 in China, and to provide epidemic prevention and help with decision making. Reference [3] used the least square method of SEIR partitioning and Poisson noise to estimate the basic reproduction number of COVID-19 in Japan as R0 = 2.6 (95% CI, 2.4-2.8). The experimental results show that the epidemic of COVID-19 in Japan will not end quickly, and it is ridiculous to think that COVID-19 will disappear in summer spontaneously.

The traditional epidemic model (SEIR) involves various factors and analyses, which may subject to potential bias. Therefore, it is necessary to propose a COVID-19 prediction model based on time series. Reference [4] proposed the ARIMA model that is useful to predict the spread of COVID-19, and then continuously improved the model by updating the data set. The experimental results show that it has good consistency with the actual epidemic spread.

Based on the ARIMA model, we perform the simply model on the epidemic data from February 24 to March 30, 2020 in Italy and then predict the epidemiological trend of COVID-19 in the next 5 days. Table 1 records the source of the original data and the description of the data set.

**Table 1.** Specifications Table.

|  |  |
| --- | --- |
| Subject | Brief analysis of COVID-2019 in Italy |
| How data were acquired | http://www.nhc.gov.cn/xcs/xxgzbd/gzbd_index.shtml<br>https://news.sina.cn/zt_d/yiqing0121 |
| Data Format | Data have been analyzed. An Excel file with data has been Uploaded |
| Dataset description | Descriptive analysis of the data, including the cumulative number of diagnoses and the difference between the number of diagnoses on the day and the number of diagnoses on the previous day<br>$(X_n - X_{n-1})$ |
| Data value | The data are reliable and represent a true epidemic situation |

## 2. Data description

The data used in this paper are sourced from the statistics of the National Health Commission(http://www.nhc.gov.cn/xcs/xxgzbd/gzbd_index.shtml) and then compiled by the website (https://news.sina.cn/zt_d/yiqing0121). Here, the cumulative number of confirmed diagnoses, new diagnoses, deaths, and cures were counted in Italy for 36 days from February 24, 2020 to March 30, 2020. A time series database was established using Excel 2019 ^[5]^. We apply the ARIMA model to predict the cumulative number of diagnoses and the number of newly diagnosed patients ^[6]^.

## 3. Models

### 3.1. Materials

ARIMA models include Autoregressive models (AR), Moving Average models (MA), Autoregressive Moving Average models (ARMA), and Autoregressive Integrated Moving Average model (ARIMA) ^[7]^.

The basic model expression of ARIMA (p, d, q) is:

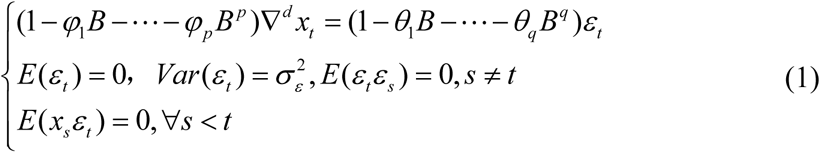

The Augmented Dickey-Fuller (ADF) unit-root test helps in estimating whether the time series is stationary ^[8]^. Log transformation and differences are the preferred approaches to stabilize the time series. Wolds decomposition theorem and Cramers decomposition theorem construct the theoretical basis of ARIMA model fitting stationary sequences ^[6]^.

Parameters of the ARIMA model were estimated by autocorrelation function (ACF) graph and partial autocorrelation (PACF) correlogram. We use R to statistically analyze the fitted predictions of the cumulative number of confirmed and newly diagnosed COVID-19 in Italy, and the significance level is set at *α =*0.05 ^[9]^.

Steps:

1. Establish the observed time series database;
2. Check the stationarity of the observation data. If the sequence is not stationary, perform a difference or logarithmic transformation until it becomes a stationary time series;
3. Calculate the ACF and PACF of the stationary sequence, and use ARIMA model to identify preliminary values of the autoregressive order, p, the order of differencing, d, and the moving average order, q.
4. Perform model tests, including the significance test of the model and the significance test of the parameters.
5. To predict the epidemic situation in the next 5 days.

### 3.2. Parameters and tests

This paper counts the epidemic situation in Italy from February 24, 2020 to March 30, 2020, and the time span is 36 days. Before modeling, we analyze the original sequence to see whether it has specific trend. The original sequence is shown in Figure 1. We can clearly see that the sequence is non-stationary because it shows a clear upward trend. In view of ARIMA modeling requires a stationary sequence, it is necessary to perform a difference or logarithmic transformation on the sequence.

**Figure 1.**
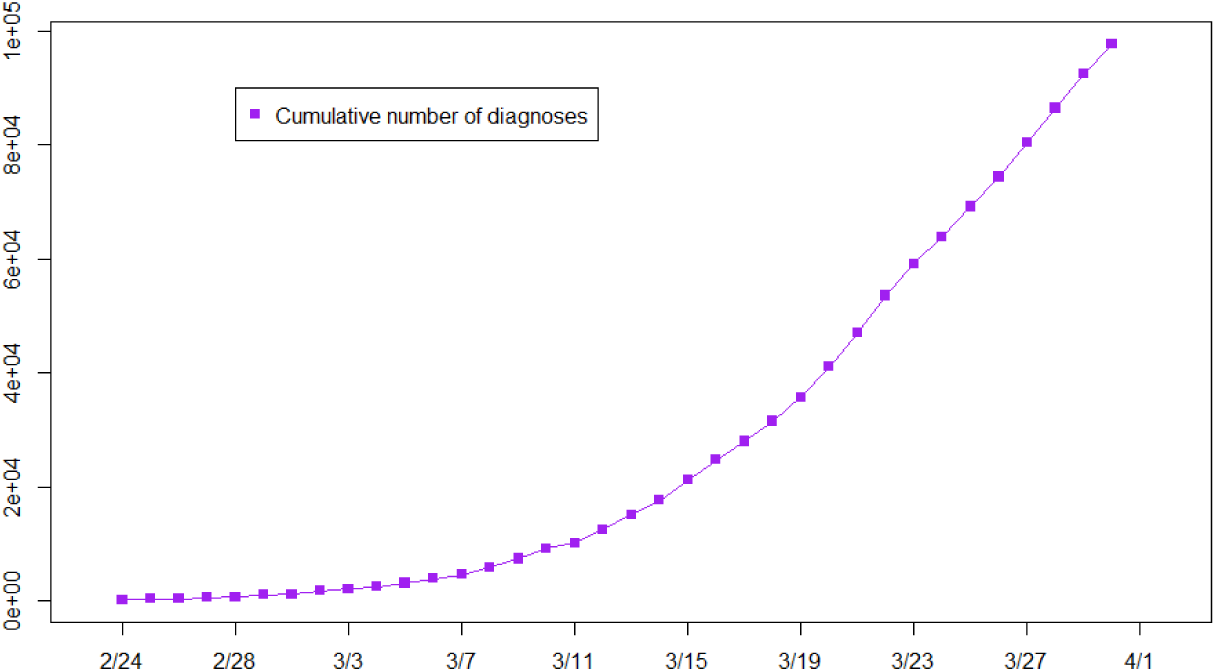
cumulative diagnoses in Italy.

Although the second-order difference of the sequence shows stationarity. We do want to mention, however, that overfitting can be used as a diagnostic tool, which will lose important information of the original sequence, it is not always the case that more is better. Overfitting leads to less-precise estimators, and adding more parameters may fit the data better but may also lead to bad forecasts. So, in this paper we first perform a logarithmic transformation on the cumulative confirmed original sequence, and then performs a first-order difference based on the logarithmic sequence. The differential logarithmic sequence is shown in Figure 2.

**Figure 2.**
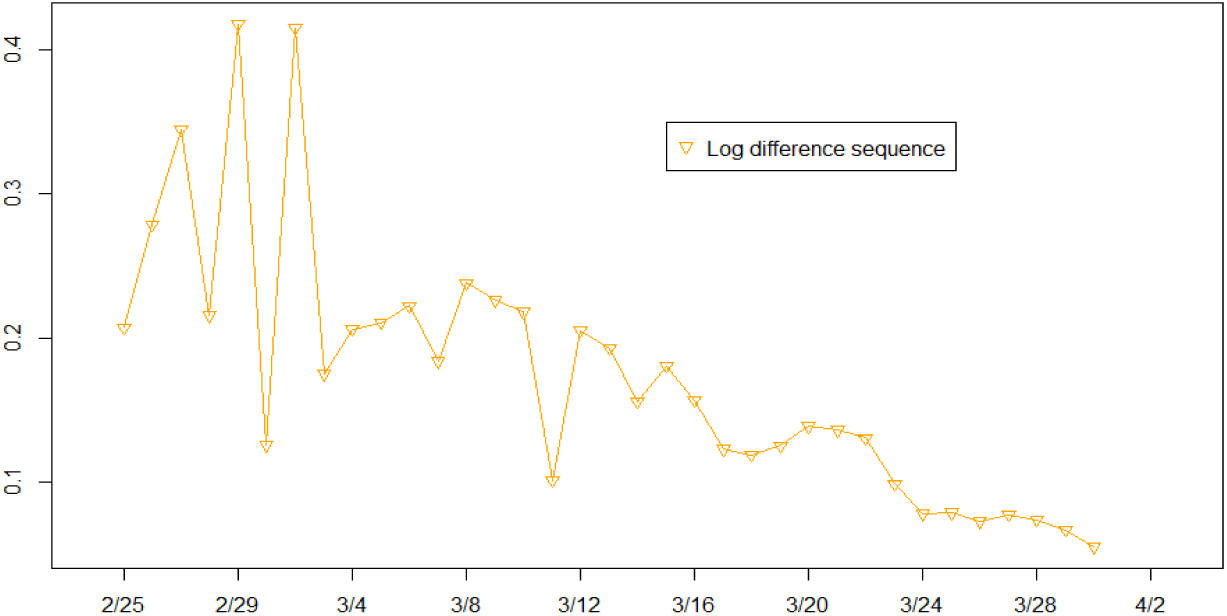
Logarithmic difference sequence of cumulative diagnoses.

R statistical software was used to calculate the ACF and PACF of a logarithmic difference sequence, the results are as follows:

The ARIMA (2,1,0) model was constructed to fit the logarithmic sequence of cumulative diagnoses, and the residual sequence after fitting was tested to be a white noise sequence, that is, the model was significant. The significance of the parameters was also tested. Based on this model, we predict the cumulative number of confirmed diagnoses in the next 5 days. Since it was a logarithmic sequence before, it should be converted accordingly next ^[10-11]^. Logarithmic cumulative confirmed sequence prediction is shown in Figure 4:

**Figure 3.**
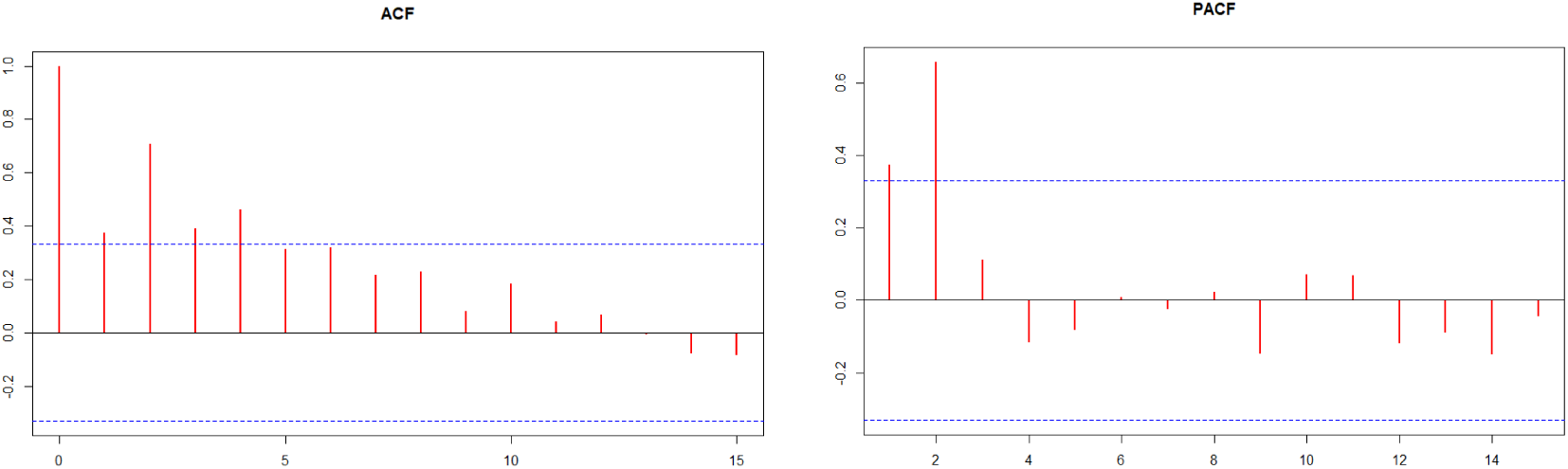
Correlogram for the cumulative logarithmic differential logarithmic.

**Figure 4.**
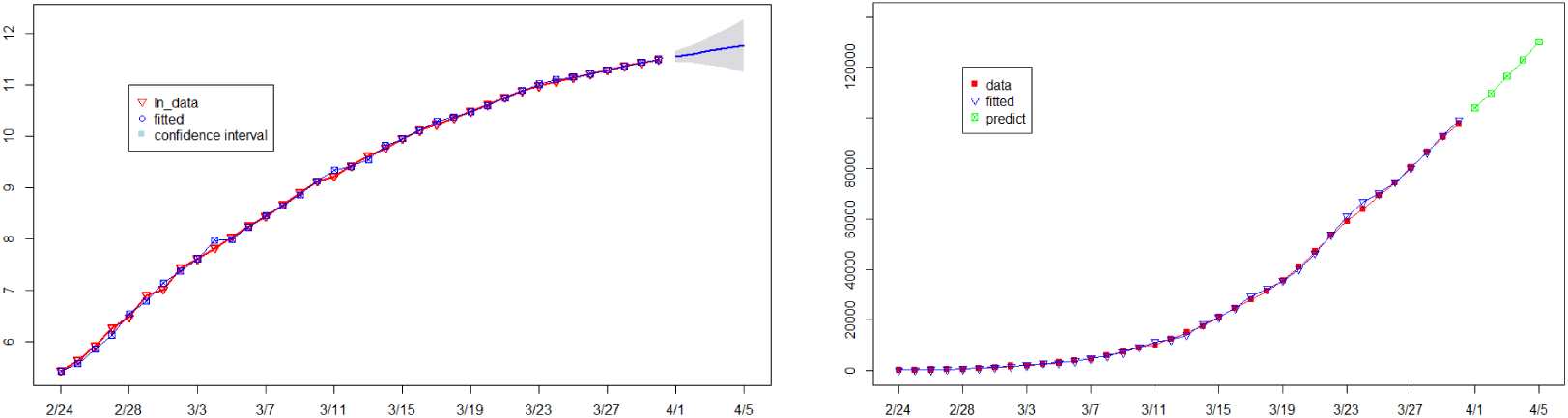
Log sequence prediction and original sequence prediction.

It is known from Figure 4: The logarithm of the cumulative number of confirmed diagnoses of COVID-2019 is in a gradual phase, that is, the growth rate of the cumulative number of confirmed diagnoses is slowing down. This is a very important signal that Italy is now at a very critical point.

The same method is used for the newly confirmed number of patients. The original sequence and the differential log sequence are as follows:

The ACF and PACF diagrams of the differential logarithmic sequence of newly diagnosed people are as follows:

For the logarithmic series of newly diagnosed patients, the ARIMA (1,1,2) model is selected to fit the predictions. The significance of the model and the significance of the parameters all pass the test.

Figure 8 shows that the difference between the number of diagnoses on the day and the number of diagnoses on the previous day is not a continuous growing process ^[12]^. The number of newly diagnosed patients has now reached a flat period, which indicates that the current prevention and control in Italy has been effective.

**Figure 5.**
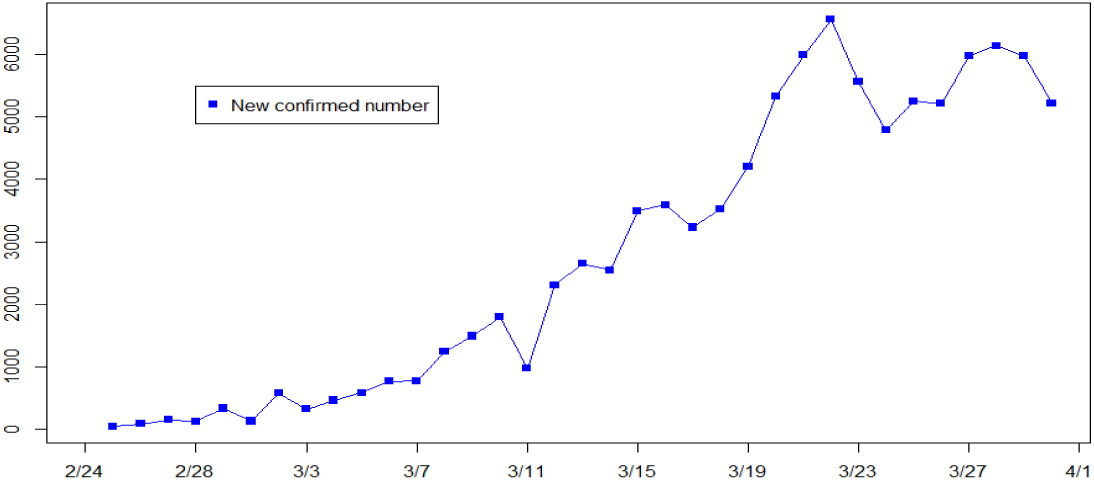
Original sequence of new confirmed diagnoses.

**Figure 6.**
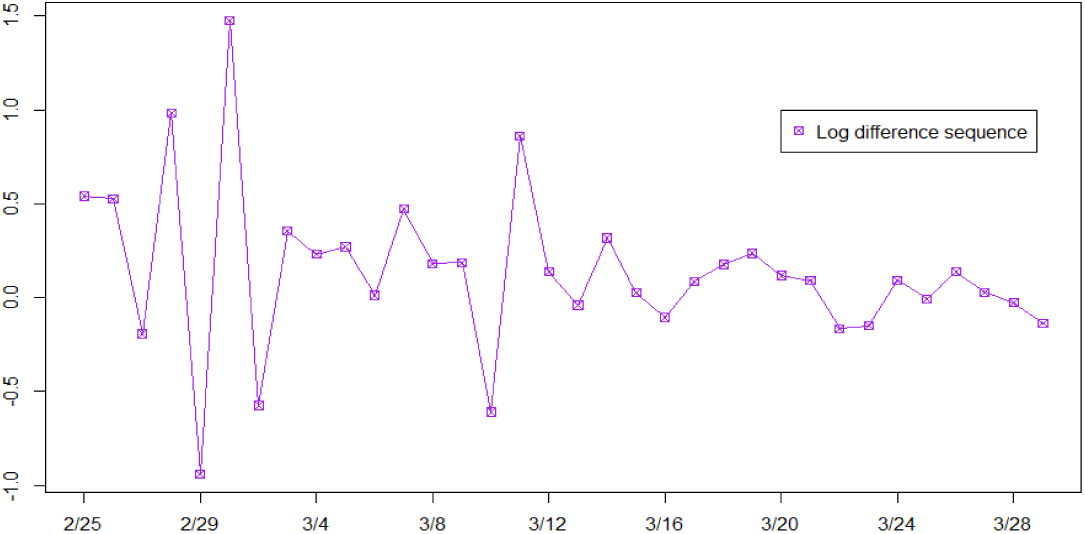
Differential logarithmic sequence of newly confirmed diagnoses.

**Figure 7.**
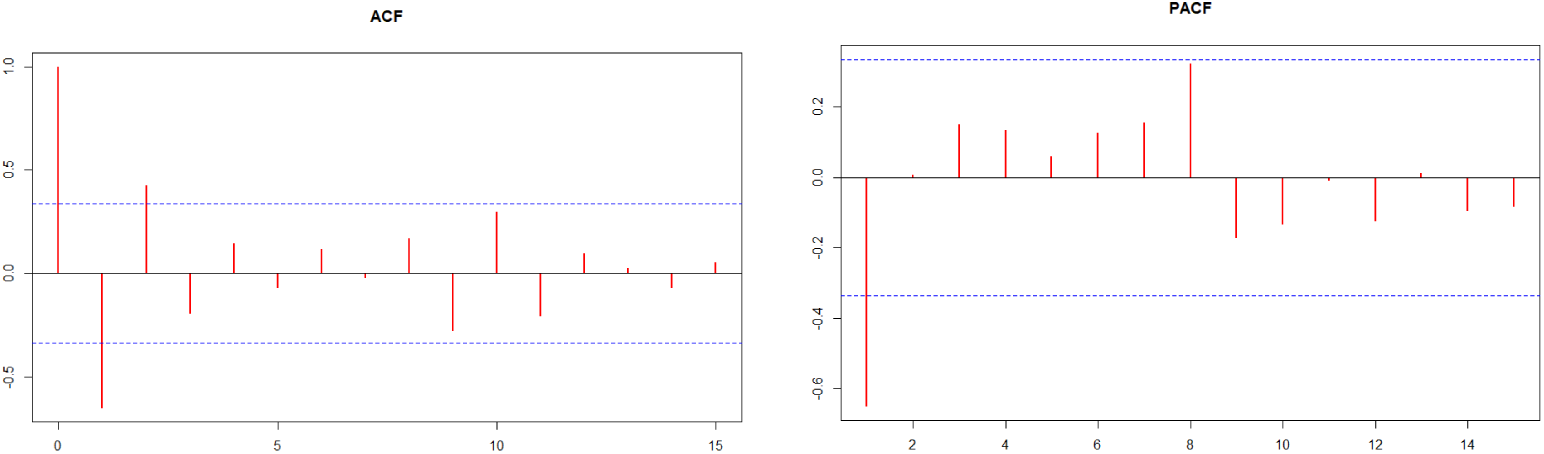
ACF and PACF diagrams of the differential logarithmic sequence of newly diagnosed persons.

**Figure 8.**
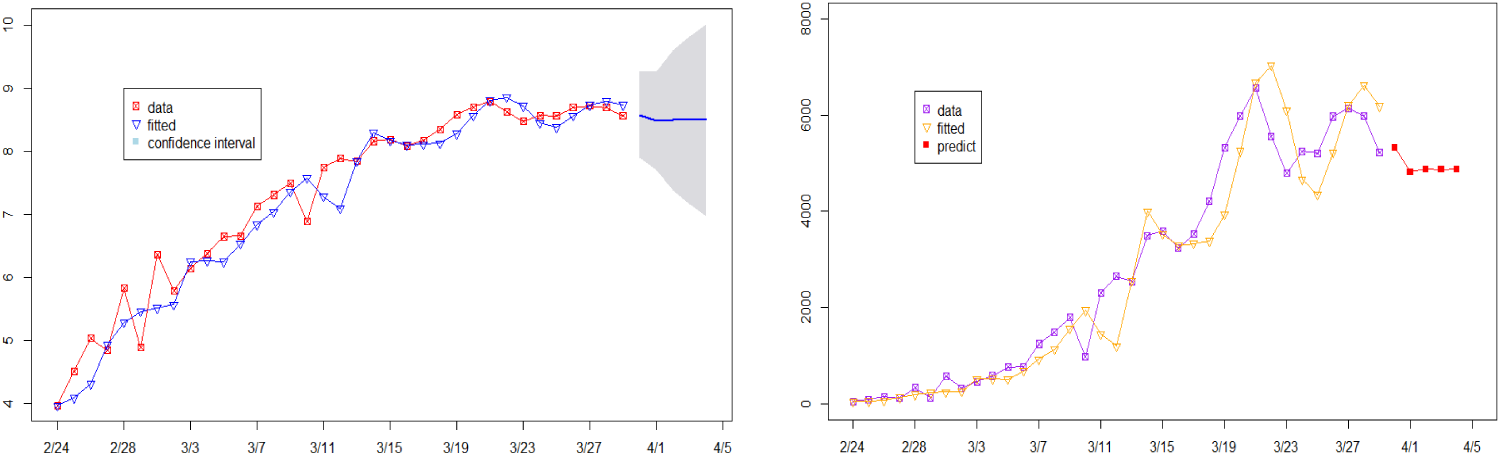
Log sequence prediction and original sequence prediction.

Based on the ARIMA model established above, the cumulative number of confirmed diagnoses and the number of newly confirmed diagnoses in the next 5 days are predicted. The 95% confidence interval data prediction is shown in Table 2.

**Table 2.** Forecast value of cumulative diagnoses and new diagnoses.

| Date | Accumulated<br>confirmed | Predictive<br>value1 | Relative<br>error1 | New<br>confirmed | Predictive<br>value2 | Relative<br>error2 |
| --- | --- | --- | --- | --- | --- | --- |
| 2020/2/24 | 230 | 229 | -0.00435 |  |  |  |
| 2020/2/25 | 283 | 265 | -0.0636 | 53 | 53 | 0 |
| 2020/2/26 | 374 | 352 | -0.05882 | 91 | 60 | -0.34066 |
| 2020/2/27 | 528 | 464 | -0.12121 | 154 | 74 | -0.51948 |
| 2020/2/28 | 655 | 701 | 0.070229 | 127 | 138 | 0.086614 |
| 2020/2/29 | 995 | 893 | -0.10251 | 340 | 197 | -0.42059 |
| 2020/3/1 | 1128 | 1275 | 0.130319 | 133 | 235 | 0.766917 |
| 2020/3/2 | 1709 | 1601 | -0.06319 | 581 | 247 | -0.57487 |
| 2020/3/3 | 2036 | 2041 | 0.002456 | 327 | 262 | -0.19878 |
| 2020/3/4 | 2502 | 2911 | 0.163469 | 466 | 521 | 0.118026 |
| 2020/3/5 | 3089 | 2983 | -0.03432 | 587 | 526 | -0.10392 |
| 2020/3/6 | 3858 | 3777 | -0.021 | 769 | 516 | -0.329 |
| 2020/3/7 | 4636 | 4745 | 0.023512 | 778 | 687 | -0.11697 |
| 2020/3/8 | 5883 | 5711 | -0.02924 | 1247 | 930 | -0.25421 |
| 2020/3/9 | 7375 | 7106 | -0.03647 | 1492 | 1139 | -0.2366 |
| 2020/3/10 | 9172 | 9273 | 0.011012 | 1797 | 1564 | -0.12966 |
| 2020/3/11 | 10149 | 11406 | 0.1238 | 1977 | 1947 | 0.01517 |
| 2020/3/12 | 12462 | 12265 | -0.01581 | 2313 | 1453 | -0.37181 |
| 2020/3/13 | 15113 | 14026 | -0.07192 | 2651 | 1200 | -0.54734 |
| 2020/3/14 | 17660 | 18404 | 0.042129 | 2547 | 2553 | 0.002356 |
| 2020/3/15 | 21157 | 21147 | -0.00047 | 3497 | 3999 | 0.143552 |
| 2020/3/16 | 24747 | 24729 | -0.00073 | 3590 | 3526 | -0.01783 |
| 2020/3/17 | 27980 | 29358 | 0.049249 | 3233 | 3298 | 0.020105 |
| 2020/3/18 | 31506 | 32369 | 0.027392 | 3526 | 3331 | -0.0553 |
| 2020/3/19 | 35713 | 35468 | -0.00686 | 4207 | 3382 | -0.1961 |
| 2020/3/20 | 41035 | 40127 | -0.02213 | 5322 | 3934 | -0.2608 |
| 2020/3/21 | 47021 | 46467 | -0.01178 | 5986 | 5253 | -0.12245 |
| 2020/3/22 | 53578 | 53784 | 0.003845 | 6557 | 6676 | 0.018149 |
| 2020/3/23 | 59138 | 61087 | 0.032957 | 5560 | 7024 | 0.263309 |
| 2020/3/24 | 63927 | 66724 | 0.043753 | 4789 | 6102 | 0.27417 |
| 2020/3/25 | 69176 | 70079 | 0.013054 | 5249 | 4661 | -0.11202 |
| 2020/3/26 | 74386 | 74625 | 0.003213 | 5210 | 4343 | -0.16641 |
| 2020/3/27 | 80359 | 80214 | -0.0018 | 5973 | 5225 | -0.12523 |
| 2020/3/28 | 86498 | 86307 | -0.00221 | 6139 | 6206 | 0.010914 |
| 2020/3/29 | 92472 | 93171 | 0.007559 | 5974 | 6622 | 0.10847 |
| 2020/3/30 | 97689 | 99195 | 0.015416 | 5217 | 6180 | 0.184589 |
| 2020/3/31 |  | 103997 |  |  | 5316 |  |
| 2020/4/1 |  | 109852 |  |  | 4822 |  |
| 2020/4/2 |  | 116560 |  |  | 4879 |  |
| 2020/4/3 |  | 123034 |  |  | 4872 |  |
| 2020/4/4 |  | 130194 |  |  | 4873 |  |

## 4. Conclusions and recommendations

R statistical software was used to construct a trend-fitting forecast based on the ARIMA model. The National Health Commission and statistical data are used to fit this epidemic trend. It can be seen that the epidemic will continue for some time. From the perspective of statistical analysis, although we need more data to make more detailed predictions, in fact, the number of confirmed new coronaviruses in Italy is still increasing, and effective prevention and control measures are still needed.

The growth rate of newly diagnosed patients in Italy has slowed down. And it is expected to reach the inflection point in early April. Before the turning point of the epidemic comes, we must not relax our vigilance and continue to carry out various government-prevented measures strictly, such as strengthening the work of regional isolation, doing effective personal protection, and organizing existing medical forces for rapid treatment.

As for the government, the most important thing at present is to greatly reduce peoples contact and implement isolation policy. China has done very well in restrictive and preventive measures. Italy must learn from China. It is hoped that the “City Closure” decree issued by the Italian government will go in the right direction, because this is the only way to curb the spread of epidemic.

## Data Availability

URL1：http://www.nhc.gov.cn/xcs/xxgzbd/gzbd_index.shtml
URL2：https://news.sina.cn/zt_d/yiqing0121

https://news.sina.cn/zt_d/yiqing0121

## Notes

### Competing Interest Statement

The authors have declared no competing interest.

### Funding Statement

No funding

